# Causal effects of maternal circulating amino acids on offspring birthweight: a Mendelian randomisation study

**DOI:** 10.1101/2022.04.15.22273911

**Authors:** Jian Zhao, Isobel D Stewart, Denis Baird, Dan Mason, John Wright, Jie Zheng, Tom R Gaunt, David M Evans, Rachel M Freathy, Claudia Langenberg, Nicole M Warrington, Deborah A Lawlor, Maria Carolina Borges, the MR-PREG collaboration

**Author notes:** Equal senior authors (Deborah A Lawlor, Maria Carolina Borges). **Author contributions:** DAL and MCB developed the idea for the project, JZ, DAL, MCB designed the study and analysis plan, JZ undertook all analyses, IDS, CL provided additional amino acid summary data that was not in the public domain at the time of starting analyses, DAL, DM, JW obtained and managed genomic and phenotypic data collection in Born in Bradford. JZ wrote the first draft of the paper with input from DAL and MCB and all authors contributed to interpretation of results and critical revision of drafts.

## Abstract

Amino acids are key to protein synthesis, energy metabolism, cell signaling and gene expression; however, the contribution of specific maternal amino acids to fetal growth is unclear. We explored the effect of maternal circulating amino acids on fetal growth, proxied by birthweight, using two-sample Mendelian randomization and summary data from a genome-wide association study (GWAS) of serum amino acids levels (sample 1, n = 86,507) and a maternal GWAS of offspring birthweight, adjusting for fetal genotype effects (sample 2, n = 406,063 with maternal and/or fetal genotype effect estimates). A total of 106 independent single nucleotide polymorphisms (SNPs) robustly associated with 19 amino acids (*p* < 4.9 × 10^−10^) were used as genetic instrumental variables. Our results provide evidence that maternal circulating glutamine (59 g offspring birthweight increase per SD increase in maternal amino acid level, 95% CI: 7, 110) and serine (27 g, 95% CI: 9, 46) raise, while leucine (−59 g, 95% CI: -106, -11) and phenylalanine (−25 g, 95% CI: -47, -4) lower offspring birthweight. Our findings strengthen evidence for key roles of maternal circulating amino acids in healthy fetal growth.

## Introduction

Healthy fetal growth and development are essential for survival, short-term, and potentially longer-term health^1-3^. Amino acids are essential for the synthesis of protein and numerous other molecules, as well as for the modulation of multiple cell signalling pathways. It has been estimated that amino acids need to be supplied at rates between 10 and 60 g/ day per kg fetus for adequate fetal growth^4^. In vivo human studies highlight complex interactions between maternal, placental and fetal mechanisms in how different amino acids are delivered to the fetus^5-7^. However, evidence from human studies on how maternal amino acids influence fetal growth and development is scarce, with inconsistent conclusions from small studies of amino acids supplementation in pregnancies at risk of fetal growth restriction^8-11^.

Mendelian randomisation (MR) is an approach where genetic variants, robustly associated with a modifiable exposure, are used as instrumental variables to infer the causal relationship between the exposure and an outcome of interest^12,13^. Maternal genetic variants, normally single nucleotide polymorphisms (SNPs), have been increasingly used as instrumental variables to examine the causal relationships between genetically influenced intrauterine exposures and offspring birthweight in MR analysis^14-17^. These have confirmed the causal effect of maternal smoking during pregnancy on slower fetal growth as assessed by repeated ultrasound scan and lower birthweight^18,19^, and of pre-pregnancy BMI and higher fasting glucose on higher birthweight^17^. MR is less likely to be biased by the socioeconomic, environmental, behavioural and health factors that confound conventional multivariable analyses, although it is subject to other sources of bias such as weak instrument bias and bias from unbalanced horizontal pleiotropy (discussed below)^20-23^. It may also provide a long-term (possibly across the whole life course) assessment of between person differences in an exposure. In relation to this study, it could establish evidence of the effect of maternal circulating amino acids during pregnancy on fetal growth (as indicated by birthweight).

The aim of this study was to use two-sample MR to estimate the potential causal effect of maternal serum levels of the 20 established amino acids on offspring birthweight in up to 406,063 individuals with maternal and/or fetal genotype effect estimates.

## Results

### Pair-wise genetic correlations across amino acids

Using data from up to 86,507 individuals, we selected 112 SNPs strongly (*p* < 4.9 × 10^−10^) and independently (r^2^ < 0.05 and ≥ 1 Mb on each side of the sentinel SNP) associated with the blood concentration of at least one of the 20 amino acids (Supplementary Table 1). Using a list of 110 independent genetic variants after more stringent LD-clumping (r^2^ < 0.01 within a 10,000 kb window), we calculated the pair-wise correlation coefficients for the SNP-amino acid effect estimates based on summary data from previously published genome-wide association studies (GWAS)^24,25^. We were able to extract 108 SNP-amino acid effect estimates across 8 Nuclear Magnetic Resonance (NMR) spectroscopy measured amino acid concentrations (alanine, glutamine, histidine, isoleucine, leucine, phenylalanine, tyrosine and valine) from the GWAS conducted by Kettunen, et al. ^24^ and 72 SNP-amino acid effect estimates across 7 Mass Spectrometry (MS) measured amino acids (asparagine, glycine, lysine, methionine, proline, serine and tryptophan) from the GWAS conducted by Shin, et al. 25.

We calculated pair-wise genetic correlations between the amino acids on each measurement platform, with the results presented in Fig. 1. Branched chain amino acids (BCAAs; valine, leucine and isoleucine) were highly correlated with one another (r = 0.77 to 0.91) and serine and glycine were also strongly correlated (r = 0.65). These results point to the clustered nature of genetic regulation of circulating amino acids, which is likely reflecting shared metabolic pathways between amino acids. As an example, valine, leucine and isoleucine are metabolized by a series of reactions catalyzed by the same enzymes to generate intermediates for the citric acid cycle (also known as the tricarboxylic acid cycle or the Krebs cycle) (Supplementary Fig. 1). Therefore, higher/lower activity of this pathway will affect the concentration of the three branched-chain amino acids. Likewise, serine and glycine are intertwined due to their interconversion as part of one-carbon metabolism, which is essential for nucleotide synthesis and methylation reactions involved in epigenetic regulation and serine is intensively involved in glycine biosynthesis within the glycine metabolism pathway (Supplementary Fig. 2).

**Fig. 1.**
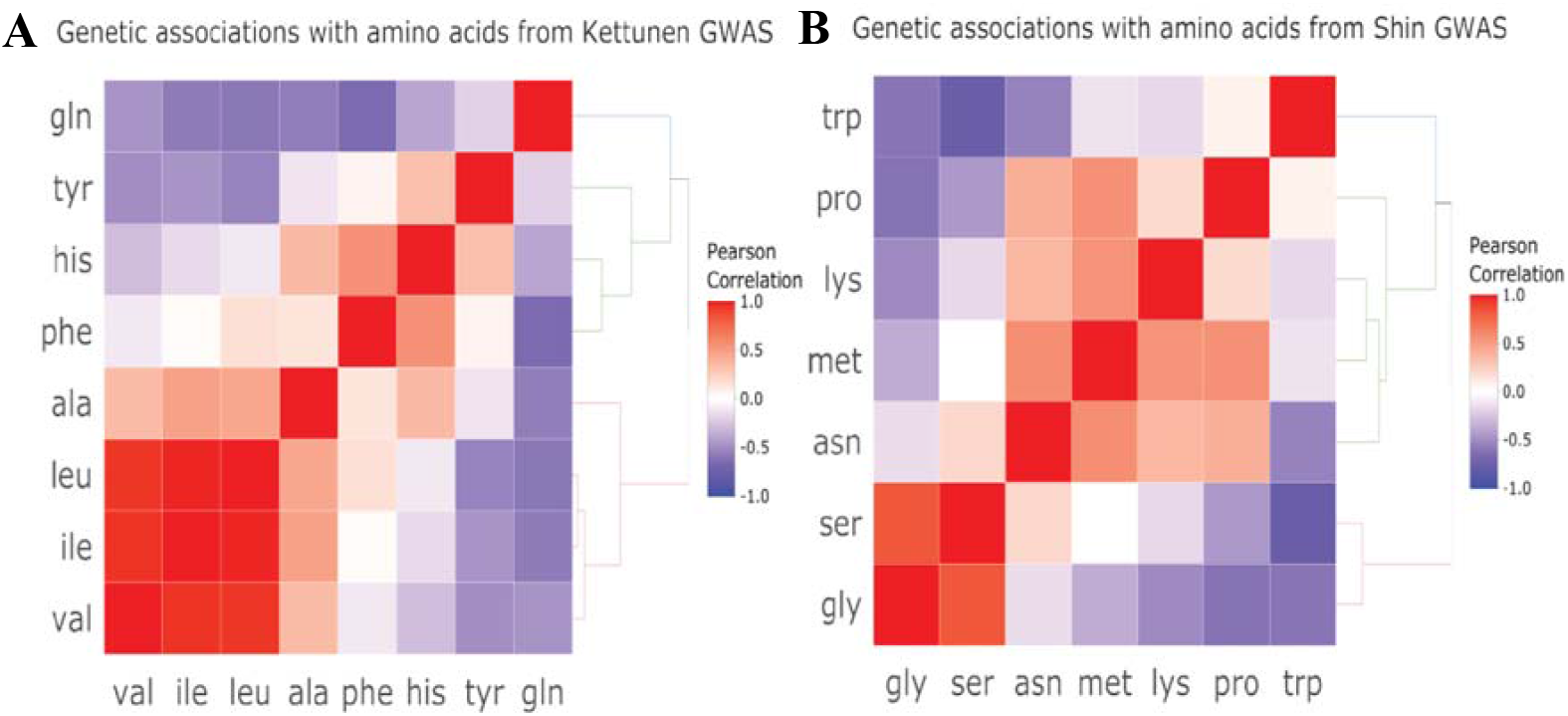
Pair-wise correlations for the SNP-amino acid effect estimates across amino acids measured using Nuclear Magnetic Resonance (NMR) spectroscopy (1A) and Mass Spectrometry (MS) (1B). Data were extracted from previously published GWAS^24,25^. Ala alanine, asn asparagine, gln glutamine, gly glycine, his histidine, ile isoleucine, leu leucine, lys lysine, met methionine, phe phenylalanine, pro proline, ser serine, trp tryptophan, tyr tyrosine, val valine.

### Genetic variants instrumenting for circulating amino acids

Five of the 110 selected genetic variants were absent from the birthweight GWAS, however we identified proxy SNPs (r^2^ ≥ 0.8) for four of them (no proxy could be found for rs142714816); this led to a total of 106 SNPs remaining as genetic instruments for the 19 amino acids after data harmonization (3 palindromic SNPs were removed and there were no available genetic variants instrumenting for cysteine) (Supplementary Table 2). These genetic variants explained a proportion of variation in circulating amino acids ranging from 0.13% (glutamate) to 4.76% (asparagine) (Supplementary Table 3).

### Estimates of causal effects of maternal amino acids on offspring birthweight

#### Main findings

In the main MR analyses, there was evidence suggesting positive causal effects of maternal serine (27 g higher offspring birthweight per SD higher serine, 95% CI: 9, 46) and glutamine (59 g, 95% CI: 7, 110) on offspring birthweight. There was also evidence of inverse causal effects of phenylalanine (−25 g, 95% CI: -47, -4) and leucine (−59 g, 95% CI: -106, -11) on offspring birthweight. There was no strong evidence for causal effects of the remaining amino acids on offspring birthweight (Fig. 2 and Supplementary Table 4). However, despite using the largest available datasets, some causal effects were imprecisely estimated and, therefore, we cannot discard the presence of biologically meaningful effects for some amino acids, such as alanine (43 g, 95% CI: -14, 100).

**Fig. 2.**
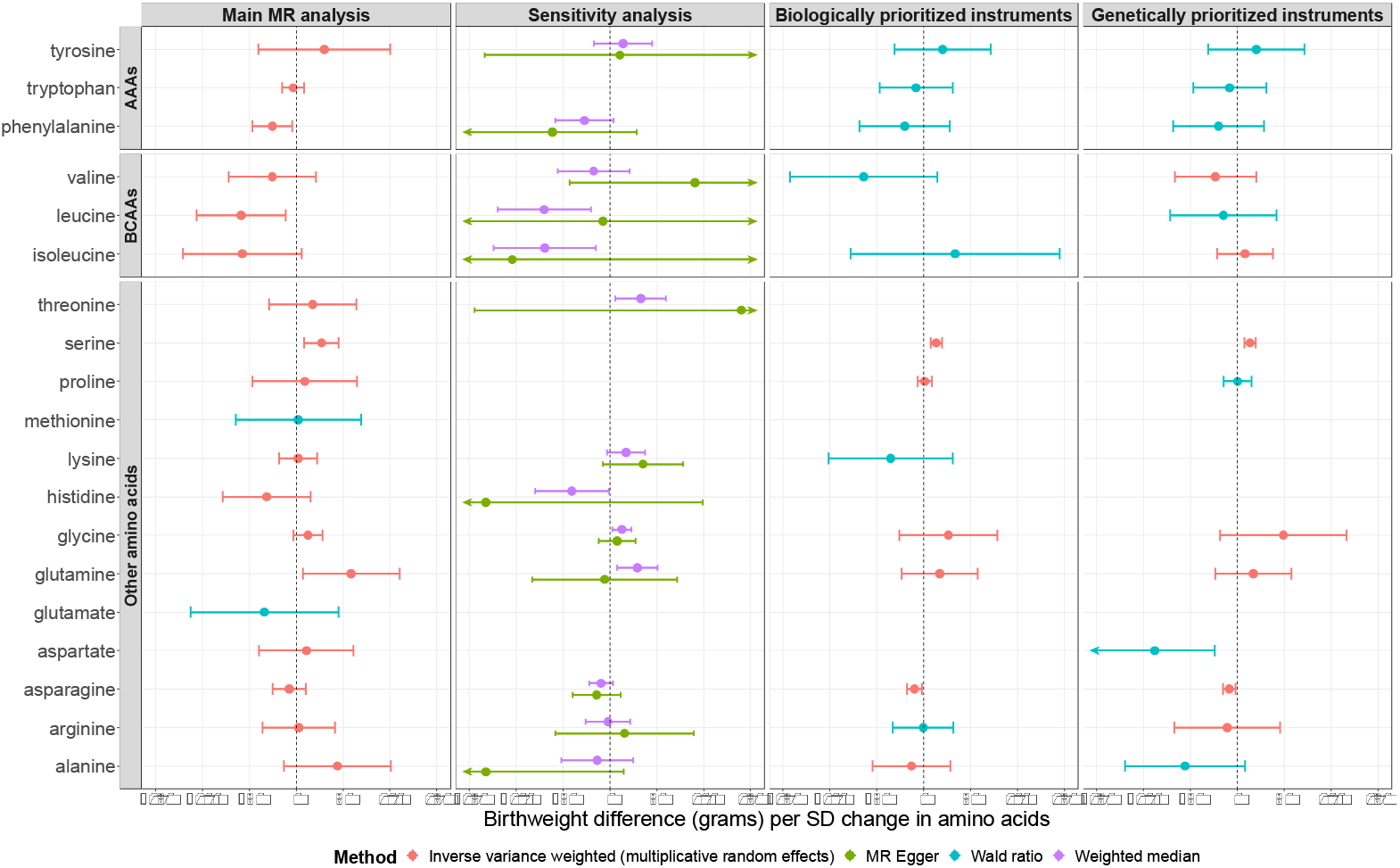
MR estimates of the effects of maternal circulating amino acids on offspring birthweight. AAAs aromatic amino acids, BCAAs branched chain amino acids. In the conservative MR analyses, we chose SNPs that were prioritized in the original metabolites GWAS^26^, where two approaches (a hypothesis-free genetic approach and a biological knowledge-based approach) were used to prioritize likely causal genes for the observed genetic associations with metabolites.

#### Sensitivity analyses to explore possible bias due to horizontal pleiotropy

##### Between SNP heterogeneity

If all SNPs instrumenting for a particular amino acid are valid (i.e., without horizontal pleiotropy effects), we would expect the effect estimates to be consistent across SNPs. To assess this, we performed leave-one-out analysis for amino acids having more than one genetic instrument and calculated Cook’s distance for amino acids having five or more genetic instruments. Cochrane’s Q statistic was used to test between SNP heterogeneity in the causal estimates.

There was evidence of between SNP heterogeneity for some of the amino acids according to the visual inspection of leave-one-out analysis results (Supplementary Fig. 3), quantification of Cook’s distance and Cochrane’s Q statistic (Table 1 and Supplementary Table 5). SNPs in or near several genes (genetically prioritized genes identified in the GWAS conducted by Lotta, et al. ^26^) that contributed to this heterogeneity were identified (Table 1). Some of these loci, such as *GCKR* (regulating glucokinase) and *CPS1* (catalysing the initial step for the urea cycle, pathway responsible for amino acids degradation and urea synthesis), have been reported to be pleiotropic in previous studies^27,28^, whereas other loci, such as *GLS2* and *PPM1K-DT*, have been found to play a key role in the regulation of glutamine and branched-chain amino acids, respectively.

**Table 1.**
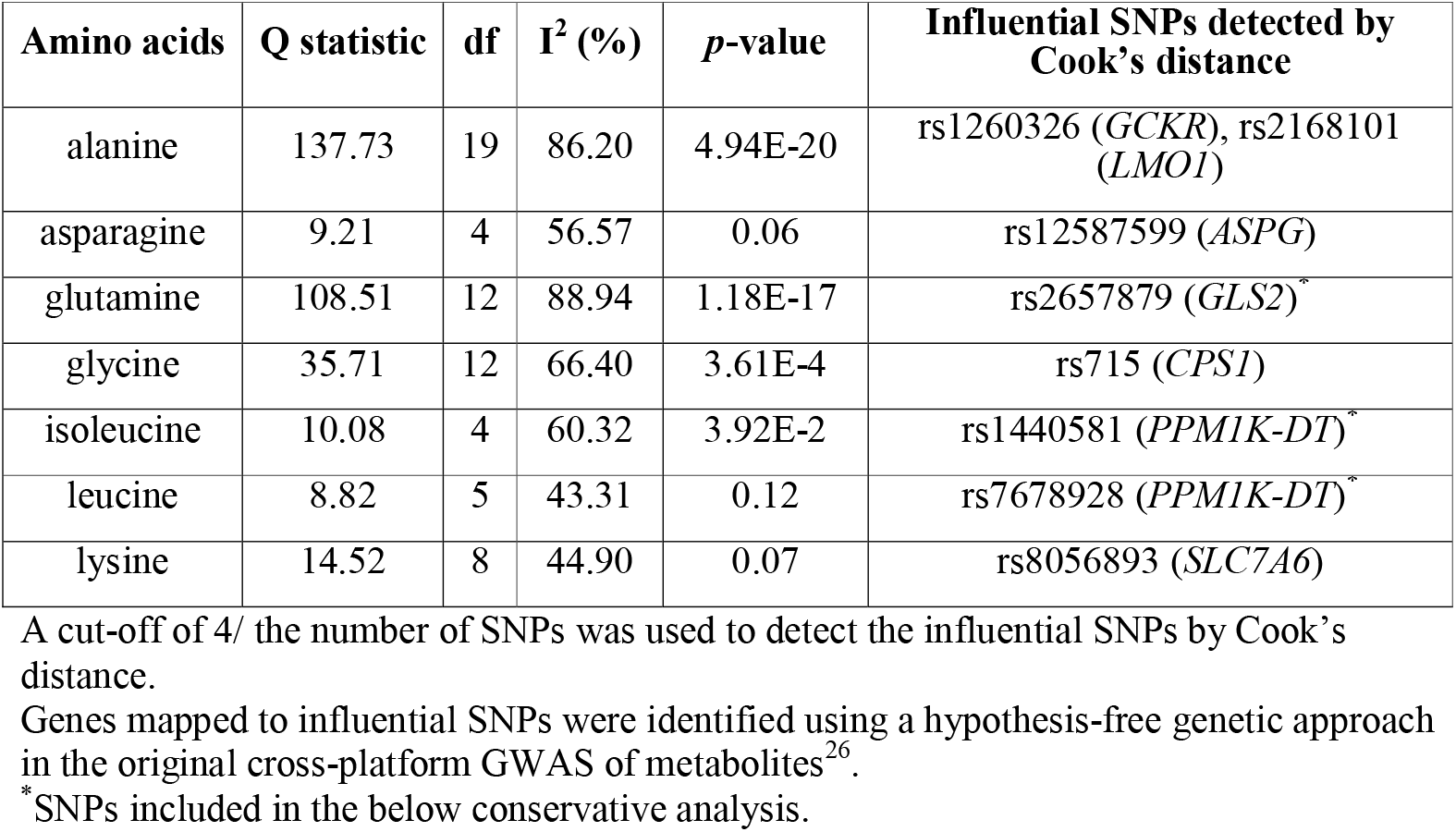
Influential SNPs detected by Cook’s distance and Cochran’s Q test results

##### MR-Egger and weighted median analyses

To explore whether these heterogeneous SNPs were causing bias in our main analyses we undertook MR sensitivity analyses that are more robust to invalid instruments (i.e., weighted median and MR-Egger regression). These require multiple SNP instruments and were conducted for 13 of the 19 amino acids with 5 or more SNPs available as genetic instruments (Fig. 2). Weighted median and MR-Egger regression analysis yielded broadly consistent results with the findings from the main IVW MR analysis (Fig. 2). The one exception was the result for alanine, which in the main IVW analysis had a positive effect on BW with wide confidence intervals including the null but in MR-Egger analysis an inverse effect size was revealed though extremely wide confidence intervals also included the null. The MR-Egger intercept also suggested that the IVW result for alanine might be biased by unbalanced horizontal pleiotropy (P=0.02) (Supplementary Table 6). For all other amino acids for which MR-Egger could be undertaken there was no strong evidence of unbalanced horizontal pleiotropy (all MR-Egger intercept *p*-values > 0.05).

##### Conservative MR analysis

In addition to these sensitivity analyses we also explored potential bias due to horizontal pleiotropy by undertaking conservative MR analyses where possible. SNPs mapped to genes directly involved in amino acids metabolism are more likely to be valid instruments for testing the effects of circulating amino acids than SNPs mapped to genes known to be highly pleiotropic (e.g., *GCKR*) or of unknown role in amino acids metabolism. Therefore, we defined conservative sets of SNPs (i.e., SNPs that are more credible instruments for circulating amino acids) and conducted MR analyses (IVW or Wald ratio) based on these. This was done by restricting the genetic instruments to those with established genetic or biological functions indicating direct causal effects on specific amino acids.

Twenty-one SNPs instrumenting for 13 amino acids were selected based on biological plausibility (selection criteria detailed in Methods section), and additional 20 SNPs instrumenting for 14 amino acids were selected based on genetic function (Supplementary Table 7). As shown in Fig. 2, compared with the findings from the main MR analysis, the conservative analysis produced broadly consistent results. A positive effect of maternal serine on offspring birthweight was confirmed for both biologically and genetically prioritized instruments. Inverse effects of asparagine and aspartate were observed in the conservative analysis, which were not consistently observed in the main MR analysis. In addition, the effect estimates for glutamine and some BCAAs (i.e., leucine and isoleucine) attenuated in the conservative analysis.

##### Analyses to explore instrument strength

The instrument strength in the main IVW MR analysis was assessed by F statistic and additionally I^2^_GX_ statistic was calculated to quantify the strength of violation of the ‘NO Measurement Error’ (NOME) assumption, which, if greater than 0.9, should not materially affect the MR-Egger regression estimates^29^. F statistic ranged from 38.74 to 7504.06 with mean F statistic across SNPs instrumenting for each amino acid ranging from 41 (glutamate) to 633 (glycine). I^2^_GX_ ranged from 0.67 to 0.99 for amino acids where MR-Egger regression analysis was performed (I^2^_GX_ < 0.9 for alanine, histidine, isoleucine, leucine and threonine and I^2^_GX_ >0.9 for the remaining amino acids) (Supplementary Table 3 for instrument strength: F statistic and I^2^_GX_), suggesting that MR-Egger analysis estimates for the former 5 amino acids should be interpreted with caution because of potential effect dilution.

##### Testing the genetic instrument relevance to maternal pregnancy amino acids

The SNPs used as genetic instruments for amino acids in this study were obtained from the largest GWAS to date that was done in non-pregnant women and men^26^. If genetic associations with amino acids differ markedly between women and men, or within women during pregnancy, our MR results could be biased. To explore this, we compared all 89 genetic associations with amino acids from the GWAS to the equivalent associations in a sample of (non-pregnant) women only (N= 4,407 Europeans from the Fenland study) and also a cohort of women (N= 2,966) with amino acids measures at 26-28 weeks gestation (the Born in Braford (BiB) cohort). A total of 67 of these genetic associations were consistent across all three samples (Fig. 3; heterogeneity P > 0.05). For the remaining 22 associations there was some evidence of heterogeneity (P ranging from 3.72E-62 to 0.04). One reason for the heterogeneity might be low imputation quality of some SNPs in BiB study, such as rs4801776 (imputation quality score INFO=0.54), rs1935 (INFO=0.65) and rs2168101 (INFO=0.69), and these SNP-amino acid pairs were depicted as extremely heterogenous in Fig. 3. Another reason for the heterogeneity is the presence of sex-specific effect between specific variants and amino acids. For example, it has been reported that there are substantial sex differences in the effect size of the variant rs715 on glycine (genetic association has higher magnitude for women than men) ^28^, and in our study it was confirmed by the magnitude difference between BiB pregnant women, Fenland non-pregnant women and GWAS general population (both pregnant and non-pregnant women higher than general population) as shown in Fig. 3. After accounting for the above potential reasons, the genetic associations with amino acids across the three data sources were broadly consistent, which provides evidence of using top hits from GWAS summary data as genetic instruments of maternal pregnancy circulating amino acids in this study. The consistency was confirmed in the comparison of the meta-analysis estimates of the genetic instrument-exposure associations across the three different data sources except for glycine, phenylalanine where substantial heterogeneity between estimates in the Fenland non-pregnant women and GWAS results in the general population was observed (Supplementary Fig. 4).

**Fig. 3.**
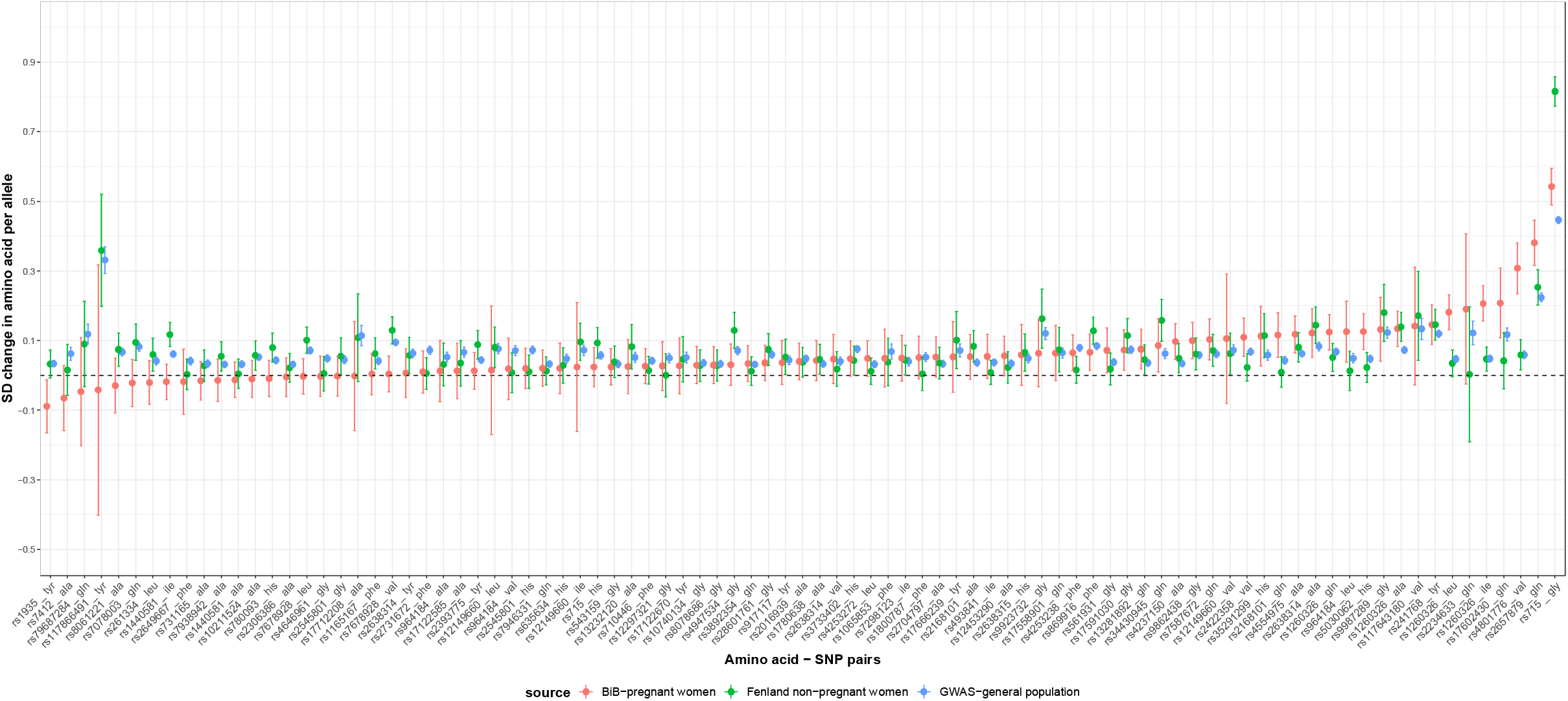
Comparisons of 89 genetic associations with amino acids between the cross-platform metabolites GWAS summary data (women and men), analysis results from the Fenland study (non-pregnant women) and the BiB study (pregnant women).

## Discussion

In the present study, we explored the causal relationships between 19 maternal circulating amino acids and offspring birthweight using two-sample MR analysis and data from the latest metabolites and birthweight GWAS, which included up to 86,507 and 406,063 participants, respectively. Our results are supportive of maternal circulating glutamine and serine having positive, and leucine and phenylalanine having negative effects on offspring birthweight.

These findings are supported by a series of sensitivity analyses exploring bias due to potential violation of MR assumptions. Despite using the largest GWAS to date for amino acids and birthweight, it should be noted that, for some amino acids, estimates were imprecise and key sensitivity analyses could not be conducted due to the low number of selected SNPs or lack of selected SNPs mapping to genes directly involved in amino acids metabolism.

Glutamine and glutamate are non-essential amino acids that become conditionally essential amino acids in catabolic stress states, which include pregnancy as fetal demand exceeds maternal synthesis^30^. Our conservative MR analyses and review of the literature suggest that the positive effect of maternal glutamine on offspring birthweight might be isoenzyme dependent^31,32^. Specifically, when glutamine was instrumented by the missense variant rs2657879 in *GLS2* there was evidence of a strong positive effect, whereas when instrumented by rs7587672, which is an eQTL for *GLS*, there was an imprecise inverse effect (Supplementary Fig. 5B). As *GLS2* encodes the enzyme which catalyzes the conversion of glutamine to glutamate and ammonia and is primarily expressed in the liver (i.e., liver-type isozyme) and *GLS* encodes the kidney-type isozyme^33^, we postulate that an overall positive causal effect of circulating glutamine level during pregnancy on offspring birthweight might be primarily driven by liver-type isoenzyme mechanism. This is indirectly supported by the positive association driven by rs17602430 (*SLC38A2*) (Supplementary Fig. 5A) that is sodium-dependent neutral amino acids (including glutamine) transporter in system A and has been reported to be functional in the supply of maternal amino acids to the fetus through the placenta^34^.

During late pregnancy glycine plays a critical role in fetal growth because it is a primary source of one-carbon necessary for both synthesis and methylation of DNA and other molecules^35^. However, there is evidence that glycine is relatively poorly transported across the human placenta and placental glycine supply is thought to be lower than fetal demand^36^. It has been hypothesized, and with some support from sheep and human pregnancy tracer studies, that maternal circulating serine is not transported to the fetal circulation via the placenta but is used within the utero-placental tissues to synthesize glycine, and via this mechanism makes an important contribution to fetal glycine supply^37,38^. Our findings appear to support this hypothesis, that is, a causal role of maternal circulating serine on offspring birthweight was found but little evidence on causal effects of maternal circulating glycine on offspring birthweight. For glycine, results from conservative analysis (Supplementary Fig. 6) based on two SNPs, rs17591030 (*GLDC*) and rs9923732 (*GCSH*), did not support a causal effect of it, though results based rs561931 (*PHGDH*) and rs4947534 (*PSPH*) suggested positive effects. The reason for this may be the distinct biological pathways involved. The *GLDC* and *GCSH* loci are involved in glycine degradation, whereas *PHGDH* and *PSPH*, encode enzymes involved in the de novo biosynthesis of serine^28^. Given the interlinked metabolism of serine and glycine, this might explain the observed potential positive effects of glycine on birthweight when estimated using *PHGDH* and *PSPH* compared to *GLDC* and *GCSH*. Further exploration of this, for example, using multivariable MR might be valuable but would require large sample sizes.

Unlike glutamine, serine and glycine, which are non-essential amino acids and use sodium-dependent A system for placental transport, BCAAs, including valine, leucine and isoleucine, use sodium-independent L system and cross the placenta much more rapidly^39^. Previous studies have reported that maternal higher concentrations of essential amino acids, including BCAAs, were associated with higher risk of intrauterine growth restricted pregnancies^7,40^. Mammalian target of rapamycin (mTOR) activity, which regulates cell growth, protein synthesis and placental amino acid transport, has been widely investigated for its role in placental function and fetal growth, and the activity of mTOR is sensitive to the supply of amino acids, particularly leucine ^41,42^. Therefore, it has been proposed that leucine works as a modulator of fetal growth through the activation of the intracellular mTOR signaling pathway^43^. In the present study, an inverse effect of maternal circulating leucine on offspring birthweight was found in the main MR analyses, which was directionally consistent with estimates from sensitivity analyses. A recent MR study revealed that higher genetically predicted BCAA levels increased the risk of type 2 diabetes^44^, and genetically elevated maternal blood glucose potentially raises offspring birthweight, which was supported by another MR study recently^17^. Thus, a positive association between BCAA levels and offspring birthweight would be expected but interestingly the findings in this study provided no evidence on this. In contrast, potentially inverse relationships between maternal circulating BCAAs and fetal growth were uncovered in this study, which was also observed in a recent metabolomic study^45^. Given the complex biological mechanisms involved in the BCAA metabolism and its close link with insulin resistance^46,47^, further MR studies in large numbers of pregnancies to dissect the impact of maternal fasting insulin and circulating BCAAs on offspring birthweight are warranted.

For phenylalanine, our main findings suggested an inverse effect on offspring birthweight, which was supported by similar effect estimates across different methods of sensitivity analyses. There is evidence, from one small observational study of 20 twins, suggesting a marked reduction in the fetal circulating concentrations of amino acids including phenylalanine transported by system L in small for gestational age twins compared to appropriate for gestational age twins, though no differences in maternal amino acids concentrations were observed between these two groups^48^, and we did not identify other studies in larger samples of the same association.

The main strengths of our study include the utilization of maternal genetic effects from the largest birthweight GWAS summary data (total sample size up to 406,063 individuals) accounting for fetal genetic effects, in a two-sample MR framework to improve causal inference. Additionally, we undertook a series of sensitivity analyses to explore bias due to violation of MR assumptions. We also used the largest and most comprehensive amino acid GWAS to select genetic instruments with validation in a sample of pregnant women. We were able to demonstrate consistent associations between the GWAS and a sample of women only, and also with an independent cohort of pregnant women, with the exception of one SNP that had previously been identified as female specific^28^. We accounted for possible population stratification by restricting to European ancestry and using GWAS summary data accounting for population structure (e.g., via principal components of ancestry or using mixed models). Our conservative analysis, conducted by restricting the genetic instruments to those with established genetic or biological functions in amino acids metabolism, minimizes the potential for bias due to horizontal pleiotropy arising from the use of SNPs that do not have specific effects on amino acids metabolism pathways. One of the limitations of our study is in one major component of the birthweight GWAS data, that is UK Biobank study, which accounts for 80% of the sample size in the birthweight GWAS, birthweight was retrospectively reported by mothers^49^. Additionally, as UK Biobank had a low response rate (∼5%) at recruitment, potential selection bias could be an issue in the genetic association studies and consequent MR analyses^50^.

In conclusion, our findings indicate that maternal genetically predicted levels of glutamine and serine increase offspring birthweight, and leucine and phenylalanine appear to reduce offspring birthweight. Despite using the largest GWAS, several causal effects were imprecisely estimated, including some that might indicate potentially important clinical effects, such as for alanine. Thus, larger GWAS of amino acids and birthweight in particular are needed to replicate our findings and elucidate mechanisms by which these amino acids could influence fetal growth. In addition, large-scale, high-quality randomized controlled trials are key to establish whether intervening on maternal circulating amino acids during pregnancy (e.g., via supplementation) can be a useful strategy to optimize healthy fetal growth.

## Methods

We selected genetic variants strongly associated with 20 different circulating amino acids from the largest GWAS available (N up to 86,507)^26^, which were validated in samples of pregnant women (N = 2,966) from the Born in Bradford study – the only study with maternal pregnancy circulating amino acids and genome wide data that we could identify^51^ – and women only (N= 4,407) using data from the Fenland study included in the main GWAS^26^. We used these genetic variants as instruments to examine the effects of maternal circulating amino acids during pregnancy on offspring birthweight in a two-sample summary data MR framework^22,52^.

### Data sources

#### Sample 1: Estimates for the association between genetic variants and amino acids

Summary data for the association between genetic variants and amino acids were retrieved from a recently conducted cross-platform GWAS of 174 metabolites that included 20 amino acids and up to 86,507 adult women and men (for individual metabolites sample sizes varied from 8,569 to 86,507)^26^. Genome-wide association analyses of up to 174 plasma metabolites were undertaken in the following cohorts of UK adults:

- Fenland study (N = 9,363 adults) in which 174 metabolites were measured using mass spectrometry (MS, Biocrates p180 kit)
- EPIC-Norfolk (N = 5,841) in which metabolites were measured using MS (Metabolon Discovery HD4)
- INTERVAL study in which metabolites were measured using MS (Metabolon Discovery HD4 platform, N = 8,455) and proton nuclear magnetic resonance (^1^H NMR, Nightingale, N = 40,905)

Given that two genotyping arrays (Affymetrix Axiom and Affymetrix SNP5.0) were used in the Fenland study, genome-wide association meta-analysis based on the two chips were first undertaken in the Fenland study and then results were meta-analysed with genome-wide association results in EPIC-Norfolk and INTERVAL for metabolites that matched those measured in the Fenland Biocrates platform. These results were further meta-analysed with publicly available GWAS summary data from two studies:

- GWAS meta-analysis of 123 metabolites measured using NMR spectroscopy on up to 24,925 individuals from 14 cohorts in Europe by Kettunen, et al. ^24^. (Data can be downloaded from: http://computationalmedicine.fi/data#NMR_GWAS)
- GWAS meta-analysis of more than 400 metabolites measured using the MS Metabolon platform on up to 7,824 individuals from two European population studies (KORA and TwinsUK) by Shin, et al. ^25^. (Data can be downloaded from: http://metabolomics.helmholtz-muenchen.de/gwas/)

A total of 20 amino acids (alanine, arginine, asparagine, aspartate, cysteine, glutamate, glutamine, glycine, histidine, isoleucine, leucine, lysine, methionine, phenylalanine, proline, serine, threonine, tryptophan, tyrosine and valine) were included in the meta-analysis. For each metabolite, a meta-analysis of z-scores was performed based on the above summary data using METAL. We calculated the SNP effect and standard error based on the z-score, sample size and minor allele frequency (MAF) reported in the abovementioned z-score based meta-analysis results using the method introduced in ref.^53^.

#### Sample 2: Estimates for the association between maternal genetic variants and offspring birthweight

Summary data for the association between genetic variants and birthweight were extracted from the most recent GWAS of birthweight which included 297,356 individuals who reported their own birthweight and 210,248 women who reported their offspring’s birthweight were combined into analysis (total n=406,063)^49^. Participants with gestational age less than 37 completed weeks (where known) or birthweight less than 2.5 kg or greater than 4.5 kg (in UK Biobank) were excluded. Associations with birthweight were in standard deviation units in this GWAS and we multiplied these by 484 (the median SD of birthweight in the 18 prospective cohorts included in this GWAS) to obtain results in grams, which are easier to interpret.

In this study, we were primarily interested in using maternal genetic variants to test the effect of maternal circulating amino acids on offspring birthweight. One challenge of utilising MR to test causal intrauterine effects on offspring outcomes, such as birthweight, is the correlation between maternal and offspring genotypes. If the offspring genetic variants affect the offspring outcome, there could be violation of the exclusion restriction assumption. The GWAS by Warrington et al. used a newly developed structural equation modelling (SEM) approach to partition maternal and fetal genetic effects on birthweight^49,54^. In this study, we used the summary data from the association of maternal genetic variants on offspring birthweight, adjusted for offspring genetic effects, as provided by the weighted linear model adjusted (WLM-adjusted) analyses, an approximation of the SEM approach.

### Genetic instrumental variable selection and harmonisation with birthweight GWAS results

A total of 112 SNPs were identified as independent signals (LD-clumping was performed using r^2^ < 0.05 and ≥ 1Mb on each side of sentinel SNP) and metabolome wide-adjusted GWAS significant with at least one of the 20 amino acids in the metabolites GWAS. In agreement with the cross-platform GWAS, we used a *p-*value < 4.9 × 10^−10^ to select SNPs strongly associated with one or more amino acids. This *p*-value threshold, which considers multiple testing resulting both from the genome-wide analyses and multiple phenotypes tested, was calculated as the conventional GWAS *p*-value threshold divided by the number of principal components explaining 95% of the variation in the 174 metabolites in the Fenland study (i.e., metabolome-wide adjusted GWAS *p*-value = 5 × 10^−8^ / 102 = 4.9 × 10^−10^).

To ensure statistical independence across SNPs instrumenting for each amino acid, we conducted a more stringent LD clumping with a cut-off of r^2^<0.01 within a 10,000 kb window, by using a 1000 Genomes European reference panel. As a result, 110 of the 112 SNPs passed the filtering and were left to be used as instruments for at least one of the 20 amino acids.

We estimated the pair-wise correlations across amino acids for the selected genetic variants to assess the potential genetic correlation between amino acids. To do that, we used the list of the above 110 selected SNPs to extract SNP-amino acid effect estimates for amino acids present in the summary data from previously published GWAS^24,25^, and then calculated and visualised the pair-wise correlation coefficients between amino acids separately for different amino acid measurement platforms (i.e., NMR and MS Metabolon).

For the 110 selected SNPs, we searched for SNP-birthweight association summary data in the GWAS of maternal genetic effects on offspring birthweight. Five SNPs (rs8061221, rs4253272, rs1065853, rs72661853 and rs142714816) were absent from the birthweight GWAS, for four of which we could identify proxy SNPs (rs7187819, rs4253282, rs7412, rs112748538) in high linkage disequilibrium (LD). No proxy SNP could be found for rs142714816, which was excluded from further analysis (Supplementary Table 8).

We harmonised the SNP-exposure and SNP-outcome association data using “ harmonise_data” function of the TwoSampleMR R package^55^. During the data harmonisation, 3 palindromic SNPs (rs28601761, rs2422358 and rs1935), with MAF of 0.42, 0.44 and 0.48, were removed because we could not unambiguously harmonise them. In the end, 106 SNPs were selected to be used as genetic instruments for the 19 amino acids (no available genetic instruments for cysteine after data harmonisation). There was considerable overlap in SNP-amino acid associations, with 6 amino acids associated with rs715, 7 amino acids associated with rs1260326 and 2 amino acids associated with other SNPs respectively as shown in Supplementary Fig. 7.

## Statistical analyses

For the main MR analysis, we used Wald ratios (for glutamate and methionine because only one SNP was available for each of the two amino acids), and multiplicative random-effect inverse variance weighted (IVW) approach^56,57^ for all other amino acids, to estimate the causal effect of maternal circulating amino acids on offspring birthweight. Both of these methods assume that all instruments are valid (e.g., no horizontal pleiotropy), even though IVW could still produce unbiased estimates in certain scenarios of IV assumptions violations (i.e., balanced horizontal pleiotropy). In this study, we focus on effect size and precision, and interpret *p-*values as evidence strength indicators^58,59^.

## MR sensitivity analyses

We conducted a series of sensitivity analysis to assess the plausibility of the core Mendelian randomization assumptions (i.e., that the SNPs are valid and strong instruments for testing the effect of maternal circulating amino acids on offspring birthweight).

### Instrument validity

Where more than one genetic instrument was available for a particular amino acid, we checked for the presence of outlier SNPs using leave-one-out analyses. For amino acids which have five or more SNPs as instrumental variables, we calculated Cook’s distance to ascertain whether any individual SNP had a disproportionate level of influence on the analysis results where a cut-off of 4/ the number of SNPs was used. We used Cochrane’s Q statistic to examine the heterogeneity between SNP-specific causal estimates. The presence of outlier SNPs and substantial heterogeneity across SNPs could be indicative of horizontal pleiotropy. Where five or more genetic instruments were available for a particular amino acid, we performed MR methods that are more robust to horizontal pleiotropy, such as weighted median^20^ and MR-Egger regression^60^. The weighted median method assumes that more than 50% of the weight in the analyses come from valid instruments and is more robust to the effects of outliers. Unlike IVW, MR-Egger does not constrain the regression slope between the SNP-amino acids and SNP-birthweight associations to go through zero. This means that the slope (MR estimate of the causal effect) is expected to be corrected in the presence of unbalanced horizontal pleiotropy as long as the INSIDE (‘Instrument Strength Independent of Direct Effect’) assumption holds. A non-zero intercept from MR-Egger indicates unbalanced pleiotropy. We compared the consistency of results across these MR methods with our main analysis results.

Furthermore, we performed a ‘conservative MR analysis’^61^, in which we selected only SNPs mapping to genes involved in amino acids metabolism pathways (e.g., amino acids biosynthesis or degradation). SNPs regulating the expression/function of these genes are more likely to be credible instruments for Mendelian randomization analysis of circulating amino acids in comparison to SNPs with an unknown role in gene regulation. We prioritised SNPs for the conservative MR analyses based on results reported in the metabolites GWAS work^26^, in which they used two approaches (a hypothesis-free genetic approach and a biological knowledge-based approach) to prioritise likely causal genes for the observed genetic associations with metabolites (details can be found in^26^). In terms of amino acids that are the focus of our study, we further looked up the list of these prioritised genes associated with amino acids in Pathway Commons database (https://www.pathwaycommons.org/) to confirm whether the functions of these genes are directly involved in the regulation of amino acids metabolism. One main selection criterion was used: a corresponding amino acid metabolism pathway can be identified in at least one data source among the many curated in that database, which reflects that the prioritized gene of a specific amino acid is directly involved in that amino acid metabolism. For those prioritised genes which were confirmed on their functional roles in the corresponding amino acids metabolism pathways, we included them in the either biologically or genetically conservative SNPs set to perform MR sensitivity analysis using Wald ratios method or multiplicative random-effect IVW method as appropriate. The results were compared with MR main analysis results.

### Instrument strength

To assess instrument strength in MR analysis, we calculated an F statistic for each genetic instrument and I^2^ GX statistic for each exposure (i.e., amino acid) in the case of MR-Egger regression^62^. A high value of I^2^_GX_ (normally greater than 0.9) would be indicative of less than 10% relative bias due to measurement error in the MR-Egger causal estimate, which is equivalent to a scenario of F-statistic greater than 10 in conventional instrumental variable analysis^62^.

### Relevance of genetic instruments

Our interest in this study is whether maternal circulating amino acids during pregnancy influence fetal growth and hence birthweight. We selected SNPs from a GWAS conducted in the general population (combining women and men) assuming that these SNPs are relevant instruments for our target population (i.e., pregnant women). We assessed the plausibility of this assumption by testing the relevance of the amino acids instruments in predicting maternal circulating amino acids during pregnancy. We were only able to identify one cohort with maternal genotype and circulating amino acids measured during pregnancy (the Born in Bradford (BiB) cohort). In BiB, amino acids were measured as part of an NMR metabolomics analysis, at 24-28 weeks of gestation (further details in^51^). Data were available for 9 of the amino acids and in 2,966 women of European ancestry. Amino acids levels were first natural log-transformed, winsorised at 5 SDs and transformed to Z scores, then adjusted for maternal age and top 10 principal components from genomic data. Each of the resulting residuals was regressed against the corresponding SNP (genetic instrument) used in the main MR analysis. A total of 89 SNP-amino acid associations were estimated from this analysis. In addition to comparing GWAS associations to the equivalent in BiB, we also compared them to the same associations in the Fenland study (non-pregnant women). In addition to comparing individual SNP associations between the GWAS, BiB and Fenland, we also compared the meta-analysis estimates of the genetic instrument-amino acid associations for each amino acid across the three different data sources.

## Data Availability

All data produced in the present work are contained in the manuscript.

## Acknowledgements

The Born in Bradford is only possible because of the enthusiasm and commitment of the Children and Parents in BiB. We are grateful to all the participants, teachers, school staff, health professionals and researchers who have made Born in Bradford happen. Data on birthweight has been contributed by the EGG Consortium using the UK Biobank Resource and has been downloaded from www.egg-consortium.org.

## References

1 Iliodromiti, S. et al. Customised and Noncustomised Birth Weight Centiles and Prediction of Stillbirth and Infant Mortality and Morbidity: A Cohort Study of 979,912 Term Singleton Pregnancies in Scotland. PLoS Med 14, e1002228, doi:10.1371/journal.pmed.1002228 (2017).

2 Huxley, R. et al. Is birth weight a risk factor for ischemic heart disease in later life? Am J Clin Nutr 85, 1244–1250, doi:10.1093/ajcn/85.5.1244 (2007).

3 Whincup, P. H. et al. Birth weight and risk of type 2 diabetes: a systematic review. JAMA 300, 2886–2897, doi:10.1001/jama.2008.886 (2008).

4 Vaughan, O., Rosario, F., Powell, T. & Jansson, T. Regulation of placental amino acid transport and fetal growth. Progress in molecular biology and translational science 145, 217–251 (2017).

5 Day, P. et al. Partitioning of glutamine synthesised by the isolated perfused human placenta between the maternal and fetal circulations. Placenta 34, 1223–1231 (2013).

6 Holm, M. B. et al. Uptake and release of amino acids in the fetal-placental unit in human pregnancies. PloS one 12, e0185760 (2017).

7 Cetin, I. et al. Maternal concentrations and fetal-maternal concentration differences of plasma amino acids in normal and intrauterine growth-restricted pregnancies. Am J Obstet Gynecol 174, 1575–1583, doi:10.1016/s0002-9378(96)70609-9 (1996).

8 Sieroszewski, P., Suzin, J. & Karowicz-Bilińska, A. Ultrasound evaluation of intrauterine growth restriction therapy by a nitric oxide donor (L-arginine). The Journal of Maternal-Fetal & Neonatal Medicine 15, 363–366 (2004).

9 Xiao, X. & Li, L. L-Arginine treatment for asymmetric fetal growth restriction. International Journal of Gynecology & Obstetrics 88, 15–18 (2005).

10 Winer, N. et al. L-Arginine treatment for severe vascular fetal intrauterine growth restriction: a randomized double-bind controlled trial. Clinical Nutrition 28, 243–248 (2009).

11 Terstappen, F. et al. Prenatal Amino Acid Supplementation to Improve Fetal Growth: A Systematic Review and Meta-Analysis. Nutrients 12, 2535 (2020).

12 Smith, G. D. & Ebrahim, S. ‘Mendelian randomization’: can genetic epidemiology contribute to understanding environmental determinants of disease? Int J Epidemiol 32, 1–22 (2003).

13 Lawlor, D. A., Harbord, R. M., Sterne, J. A., Timpson, N. & Davey Smith, G. Mendelian randomization: using genes as instruments for making causal inferences in epidemiology. Stat Med 27, 1133–1163, doi:10.1002/sim.3034 (2008).

14 Lawlor, D. et al. Using Mendelian randomization to determine causal effects of maternal pregnancy (intrauterine) exposures on offspring outcomes: Sources of bias and methods for assessing them. Wellcome Open Res 2, 11, doi:10.12688/wellcomeopenres.10567.1 (2017).

15 Lawlor, D. A. et al. Exploring the developmental overnutrition hypothesis using parental-offspring associations and FTO as an instrumental variable. PLoS Med 5, e33, doi:10.1371/journal.pmed.0050033 (2008).

16 Thompson, W. D. et al. Association of maternal circulating 25(OH)D and calcium with birth weight: A mendelian randomisation analysis. PLoS Med 16, e1002828, doi:10.1371/journal.pmed.1002828 (2019).

17 Tyrrell, J. et al. Genetic Evidence for Causal Relationships Between Maternal Obesity-Related Traits and Birth Weight. JAMA 315, 1129–1140, doi:10.1001/jama.2016.1975 (2016).

18 Brand, J. S. et al. Associations of maternal quitting, reducing, and continuing smoking during pregnancy with longitudinal fetal growth: Findings from Mendelian randomization and parental negative control studies. PLoS Med 16, e1002972, doi:10.1371/journal.pmed.1002972 (2019).

19 Tyrrell, J. et al. Genetic variation in the 15q25 nicotinic acetylcholine receptor gene cluster (CHRNA5-CHRNA3-CHRNB4) interacts with maternal self-reported smoking status during pregnancy to influence birth weight. Hum Mol Genet 21, 5344–5358, doi:10.1093/hmg/dds372 (2012).

20 Bowden, J. et al. A framework for the investigation of pleiotropy in two-sample summary data Mendelian randomization. Stat Med 36, 1783–1802, doi:10.1002/sim.7221 (2017).

21 Burgess, S., Thompson, S. G. & Collaboration, C. C. G. Avoiding bias from weak instruments in Mendelian randomization studies. Int J Epidemiol 40, 755–764, doi:10.1093/ije/dyr036 (2011).

22 Lawlor, D. A. Commentary: Two-sample Mendelian randomization: opportunities and challenges. Int J Epidemiol 45, 908–915, doi:10.1093/ije/dyw127 (2016).

23 Smith, G. D. et al. Clustered environments and randomized genes: a fundamental distinction between conventional and genetic epidemiology. PLoS Med 4, e352, doi:10.1371/journal.pmed.0040352 (2007).

24 Kettunen, J. et al. Genome-wide study for circulating metabolites identifies 62 loci and reveals novel systemic effects of LPA. Nat Commun 7, 11122, doi:10.1038/ncomms11122 (2016).

25 Shin, S. Y. et al. An atlas of genetic influences on human blood metabolites. Nat Genet 46, 543–550, doi:10.1038/ng.2982 (2014).

26 Lotta, L. A. et al. A cross-platform approach identifies genetic regulators of human metabolism and health. Nature Genetics 53, 54–64 (2021).

27 Bi, M. et al. Association of rs780094 in GCKR with metabolic traits and incident diabetes and cardiovascular disease: the ARIC Study. PLoS One 5, e11690, doi:10.1371/journal.pone.0011690 (2010).

28 Wittemans, L. B. L. et al. Assessing the causal association of glycine with risk of cardio-metabolic diseases. Nat Commun 10, 1060, doi:10.1038/s41467-019-08936-1 (2019).

29 Bowden, J. et al. Assessing the suitability of summary data for two-sample Mendelian randomization analyses using MR-Egger regression: the role of the I2 statistic. International journal of epidemiology 45, 1961–1974 (2016).

30 Neu, J. Glutamine in the fetus and critically ill low birth weight neonate: metabolism and mechanism of action. J Nutr 131, 2585S-2589S; discussion 2590S, doi:10.1093/jn/131.9.2585S (2001).

31 Foley, C. N., Mason, A. M., Kirk, P. D. W. & Burgess, S. MR-Clust: clustering of genetic variants in Mendelian randomization with similar causal estimates. Bioinformatics 37, 531–541, doi:10.1093/bioinformatics/btaa778 (2021).

32 Kutalik, Z. Commentary on: “ The contribution of tissue-specific BMI-associated gene sets to cardiometabolic disease risk: a Mendelian randomization study”. Int J Epidemiol 49, 1257–1258, doi:10.1093/ije/dyaa062 (2020).

33 Curthoys, N. P. & Watford, M. Regulation of glutaminase activity and glutamine metabolism. Annu Rev Nutr 15, 133–159, doi:10.1146/annurev.nu.15.070195.001025 (1995).

34 Huang, X. et al. Identification of placental nutrient transporters associated with intrauterine growth restriction and pre-eclampsia. BMC Genomics 19, 173, doi:10.1186/s12864-018-4518-z (2018).

35 Wang, W. et al. Glycine metabolism in animals and humans: implications for nutrition and health. Amino Acids 45, 463–477, doi:10.1007/s00726-013-1493-1 (2013).

36 Lewis, R. M. et al. L-serine uptake by human placental microvillous membrane vesicles. Placenta 28, 445–452, doi:10.1016/j.placenta.2006.06.014 (2007).

37 Hsu, J. W. et al. Unlike pregnant adult women, pregnant adolescent girls cannot maintain glycine flux during late pregnancy because of decreased synthesis from serine. Br J Nutr 115, 759–763, doi:10.1017/S0007114515005279 (2016).

38 Cetin, I. Amino acid interconversions in the fetal-placental unit: the animal model and human studies in vivo. Pediatr Res 49, 148–154, doi:10.1203/00006450-200102000-00004 (2001).

39 Battaglia, F. C. & Regnault, T. R. Placental transport and metabolism of amino acids. Placenta 22, 145–161, doi:10.1053/plac.2000.0612 (2001).

40 Paolini, C. L. et al. Placental transport of leucine, phenylalanine, glycine, and proline in intrauterine growth-restricted pregnancies. J Clin Endocrinol Metab 86, 5427–5432, doi:10.1210/jcem.86.11.8036 (2001).

41 Roos, S. et al. Mammalian target of rapamycin in the human placenta regulates leucine transport and is down-regulated in restricted fetal growth. J Physiol 582, 449–459, doi:10.1113/jphysiol.2007.129676 (2007).

42 Wullschleger, S., Loewith, R. & Hall, M. N. TOR signaling in growth and metabolism. Cell 124, 471–484, doi:10.1016/j.cell.2006.01.016 (2006).

43 Teodoro, G. F. et al. Leucine is essential for attenuating fetal growth restriction caused by a protein-restricted diet in rats. J Nutr 142, 924–930, doi:10.3945/jn.111.146266 (2012).

44 Lotta, L. A. et al. Genetic Predisposition to an Impaired Metabolism of the Branched-Chain Amino Acids and Risk of Type 2 Diabetes: A Mendelian Randomisation Analysis. PLoS Med 13, e1002179, doi:10.1371/journal.pmed.1002179 (2016).

45 Moros, G. et al. Insights into intrauterine growth restriction based on maternal and umbilical cord blood metabolomics. Sci Rep 11, 7824, doi:10.1038/s41598-021-87323-7 (2021).

46 Jang, C. et al. A branched-chain amino acid metabolite drives vascular fatty acid transport and causes insulin resistance. Nat Med 22, 421–426, doi:10.1038/nm.4057 (2016).

47 Langenberg, C. & Savage, D. B. An amino acid profile to predict diabetes? Nat Med 17, 418–420, doi:10.1038/nm0411-418 (2011).

48 Bajoria, R., Sooranna, S. R., Ward, S. & Hancock, M. Placenta as a link between amino acids, insulin-IGF axis, and low birth weight: evidence from twin studies. J Clin Endocrinol Metab 87, 308–315, doi:10.1210/jcem.87.1.8184 (2002).

49 Warrington, N. M. et al. Maternal and fetal genetic effects on birth weight and their relevance to cardio-metabolic risk factors. Nat Genet 51, 804–814, doi:10.1038/s41588-019-0403-1 (2019).

50 Munafo, M. & Smith, G. D. Biased Estimates in Mendelian Randomization Studies Conducted in Unrepresentative Samples. JAMA Cardiol 3, 181, doi:10.1001/jamacardio.2017.4279 (2018).

51 Taylor, K. et al. Differences in Pregnancy Metabolic Profiles and Their Determinants between White European and South Asian Women: Findings from the Born in Bradford Cohort. Metabolites 9, doi:10.3390/metabo9090190 (2019).

52 Evans, D. M., Moen, G. H., Hwang, L. D., Lawlor, D. A. & Warrington, N. M. Elucidating the role of maternal environmental exposures on offspring health and disease using two-sample Mendelian randomization. Int J Epidemiol 48, 861–875, doi:10.1093/ije/dyz019 (2019).

53 Zhu, Z. et al. Integration of summary data from GWAS and eQTL studies predicts complex trait gene targets. Nat Genet 48, 481–487, doi:10.1038/ng.3538 (2016).

54 Warrington, N. M., Freathy, R. M., Neale, M. C. & Evans, D. M. Using structural equation modelling to jointly estimate maternal and fetal effects on birthweight in the UK Biobank. Int J Epidemiol 47, 1229–1241, doi:10.1093/ije/dyy015 (2018).

55 Hemani, G. et al. The MR-Base platform supports systematic causal inference across the human phenome. Elife 7, doi:10.7554/eLife.34408 (2018).

56 Burgess, S., Butterworth, A. & Thompson, S. G. Mendelian randomization analysis with multiple genetic variants using summarized data. Genet Epidemiol 37, 658–665, doi:10.1002/gepi.21758 (2013).

57 Burgess, S., Small, D. S. & Thompson, S. G. A review of instrumental variable estimators for Mendelian randomization. Stat Methods Med Res 26, 2333–2355, doi:10.1177/0962280215597579 (2017).

58 Sterne, J. A. & Davey Smith, G. Sifting the evidence-what’s wrong with significance tests? BMJ 322, 226–231, doi:10.1136/bmj.322.7280.226 (2001).

59 Wasserstein, R. L. & Lazar, N. A. The ASA Statement on p-Values: Context, Process, and Purpose. The American Statistician 70, 129–133, doi:10.1080/00031305.2016.1154108 (2016).

60 Bowden, J., Davey Smith, G. & Burgess, S. Mendelian randomization with invalid instruments: effect estimation and bias detection through Egger regression. Int J Epidemiol 44, 512–525, doi:10.1093/ije/dyv080 (2015).

61 Borges, M. C. et al. Role of Adiponectin in Coronary Heart Disease Risk: A Mendelian Randomization Study. Circ Res 119, 491–499, doi:10.1161/CIRCRESAHA.116.308716 (2016).

62 Bowden, J. et al. Assessing the suitability of summary data for two-sample Mendelian randomization analyses using MR-Egger regression: the role of the I2 statistic. Int J Epidemiol 45, 1961–1974, doi:10.1093/ije/dyw220 (2016).

