## supplementary figures for "Causal effects of maternal circulating amino acids on offspring birthweight: a Mendelian randomisation study"

Supplementary information

Supplementary Fig. 1. Major metabolic pathways for branched chain amino acids degradation.

Supplementary Fig. 2. Major metabolic pathways for serine and glycine biosynthesis.

Supplementary Fig. 3. Leave-one-out analysis result

Supplementary Fig. 4. Comparison of the meta-analysis estimates of genetic instrument-exposure associations across the three different data sources (BiB study, Fenland study and metabolites GWAS in general population)

Supplementary Fig. 5. Comparison between the main MR analysis and conservative MR analysis for glutamine

Supplementary Fig. 6. Comparison between the main MR analysis and conservative MR analysis for glycine

Supplementary Fig. 7. Overlap in SNPs selected as instruments for amino acids

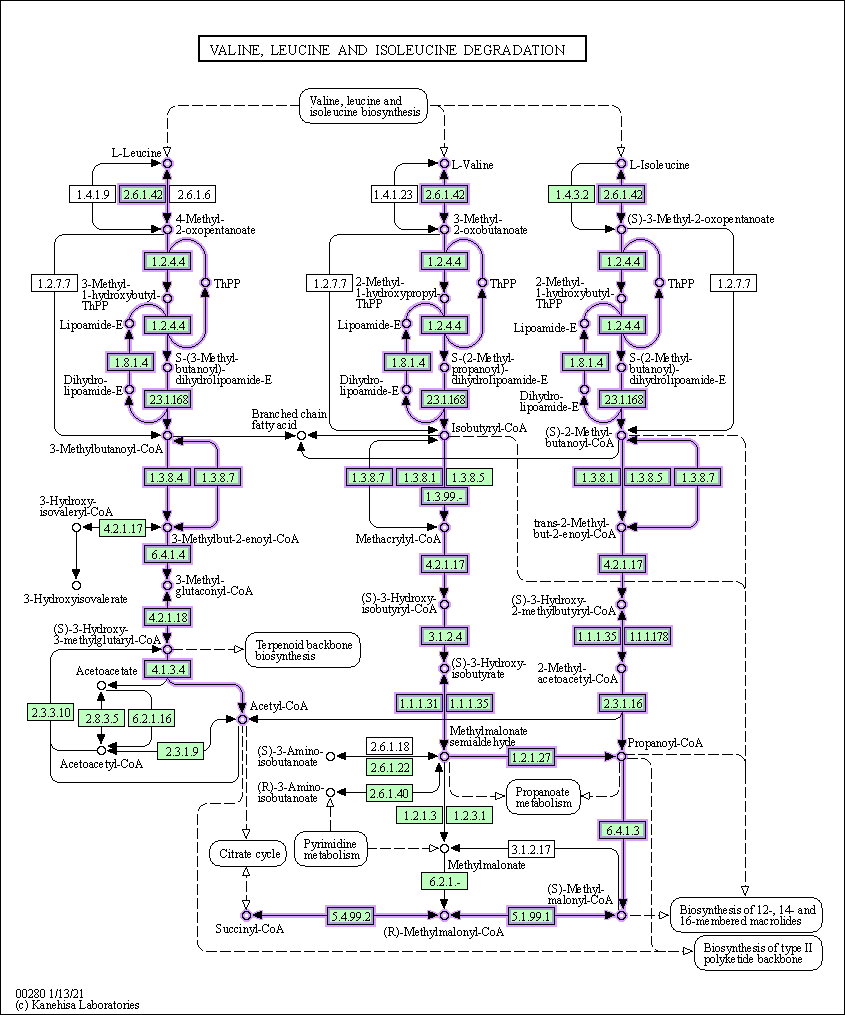

Supplementary Fig. 1. Major metabolic pathways for branched chain amino acids degradation (downloaded from KEGG Pathway Map: https://www.genome.jp/pathway/hsa00280+N00832).

Note: We will request Copyright Permission for this map from KEGG after acceptance.

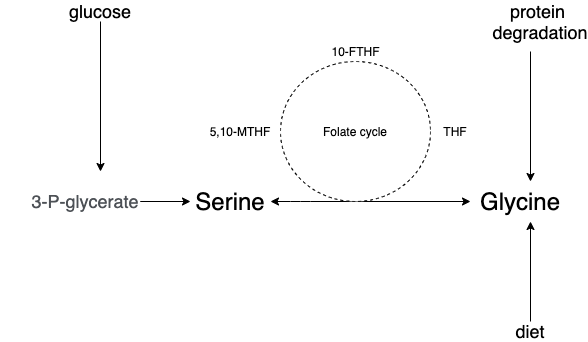

Supplementary Fig. 2. Major metabolic pathways for serine and glycine biosynthesis.

THF: tetrahydrofolate, 5,10-MTHF: N⁵-N¹⁰-methylenetetrahydrofolate, 10-FTHF: 10-formyltetrahydrofolate

Note: This is a hand-drawn pathway figure based on our knowledge for illustration purpose.

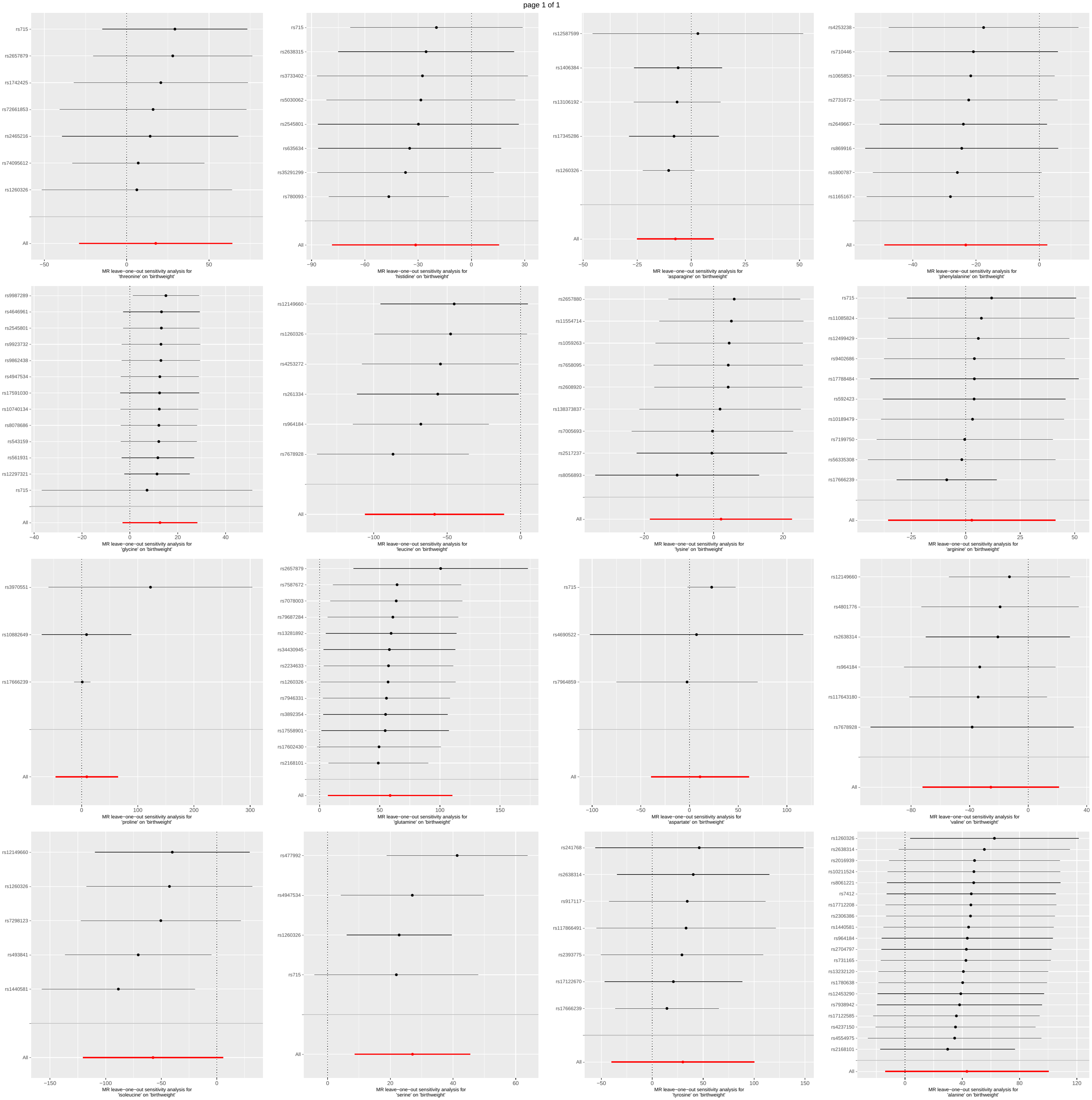

Supplementary Fig. 3. Leave-one-out analysis result

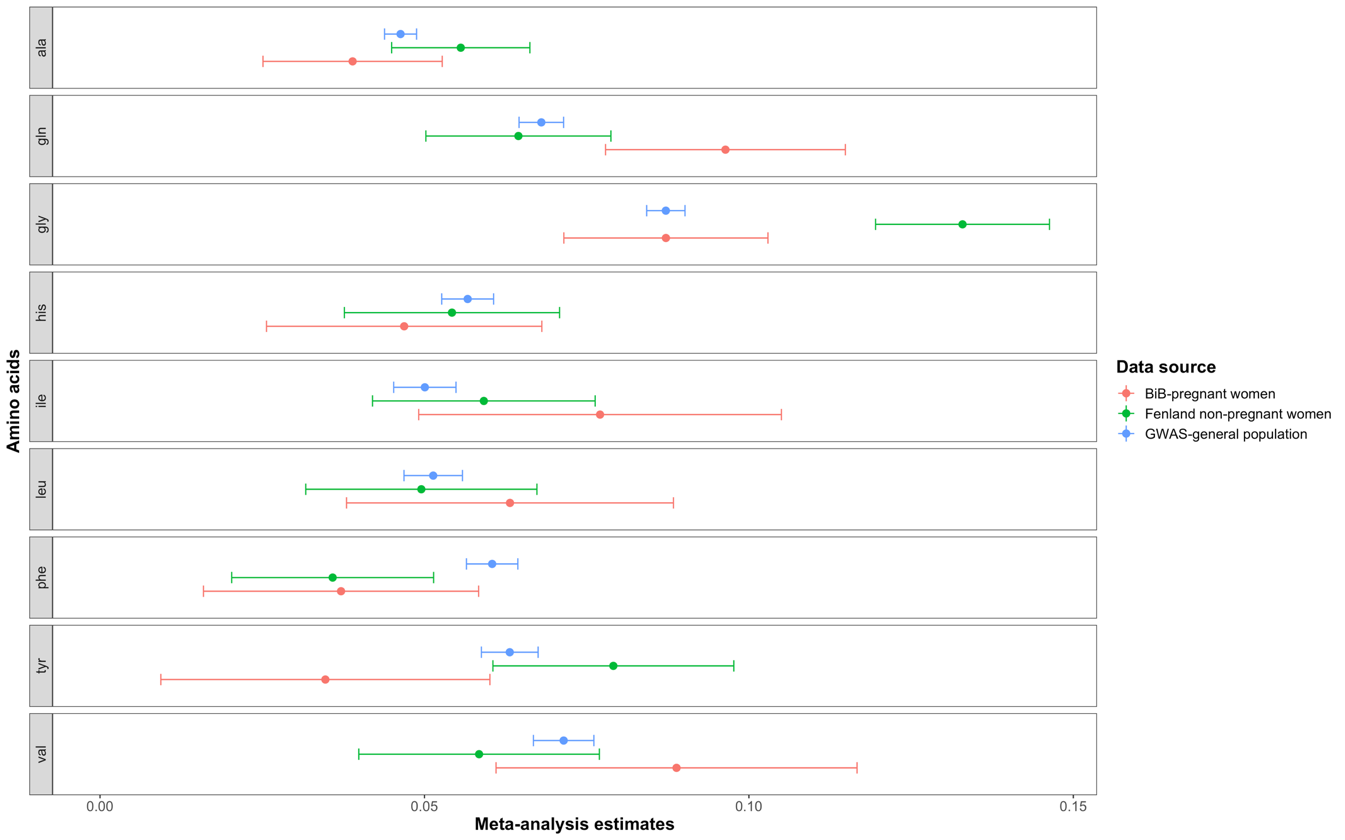

Supplementary Fig. 4. Comparison of the meta-analysis estimates of the genetic instrument-exposure associations across the three different data sources (BiB study, Fenland study and metabolites GWAS in the general population)

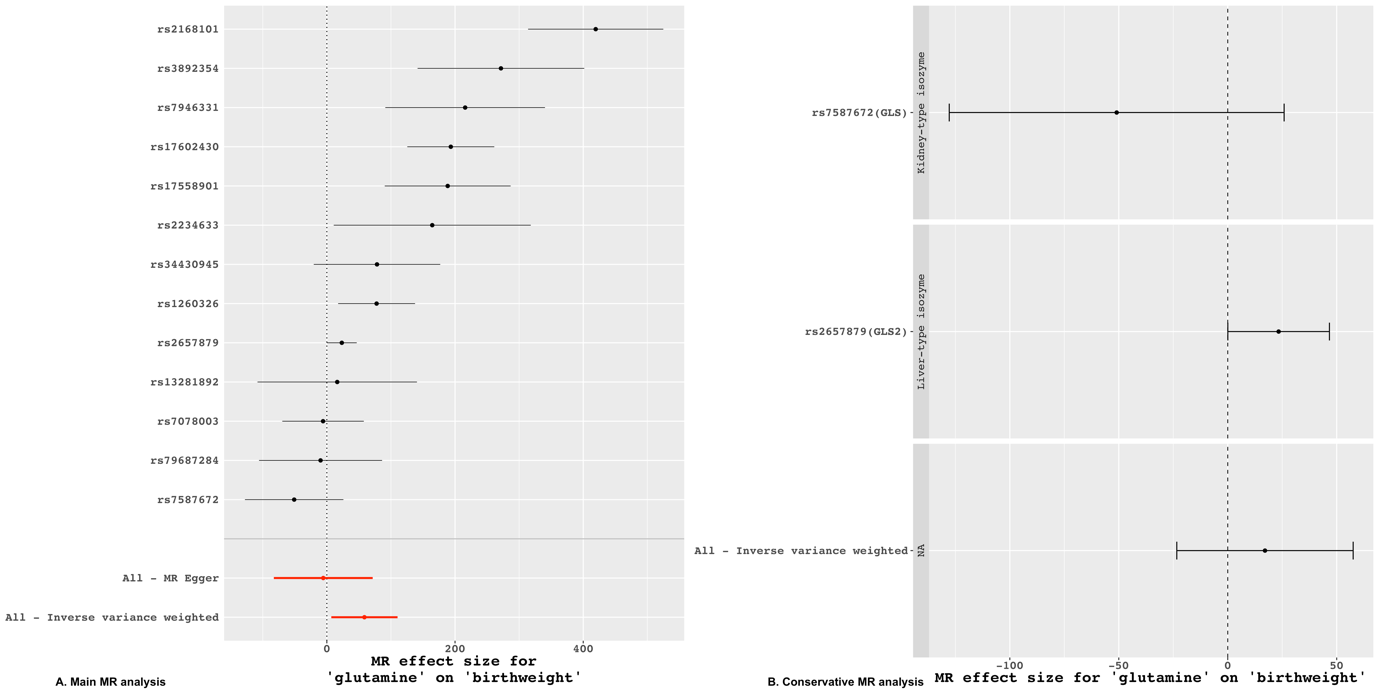

Supplementary Fig. 5. Comparison between the main MR analysis and conservative MR analysis for glutamine

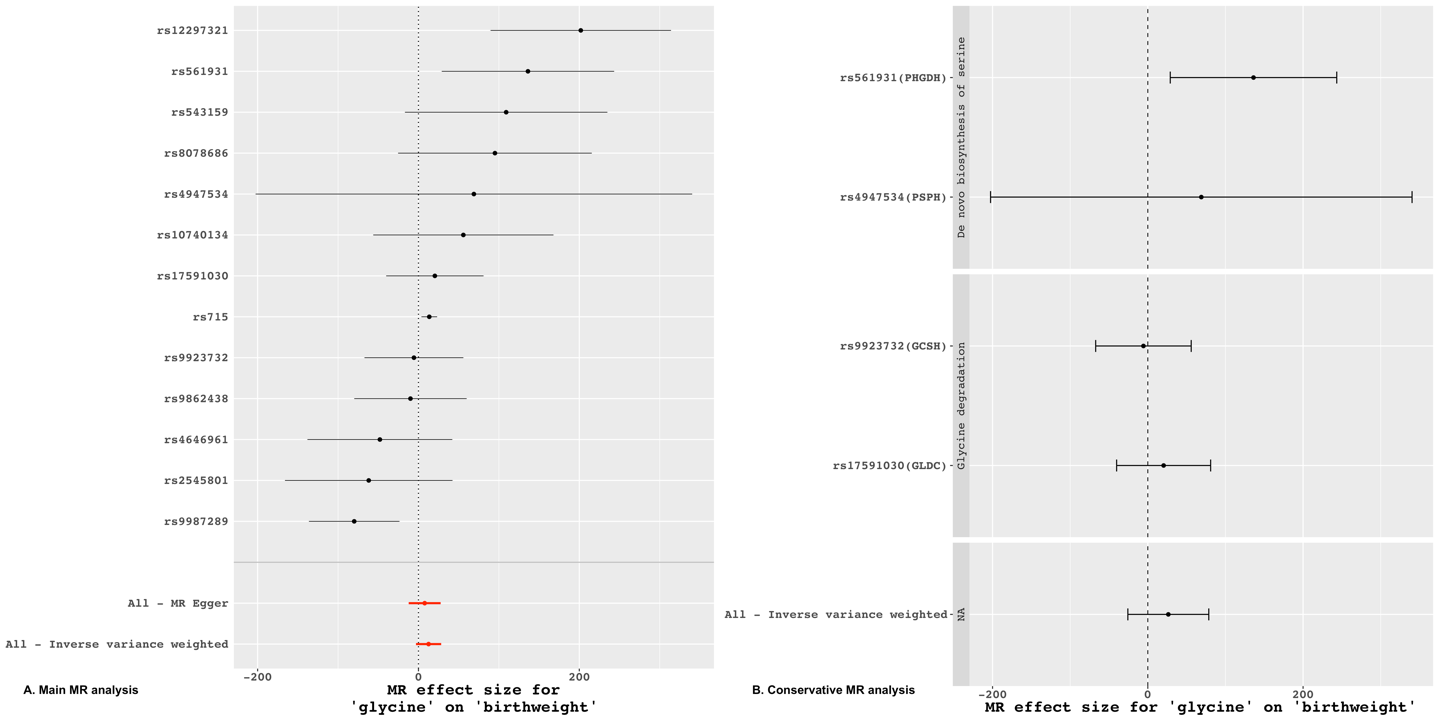

Supplementary Fig. 6. Comparison between the main MR analysis and conservative MR analysis for glycine

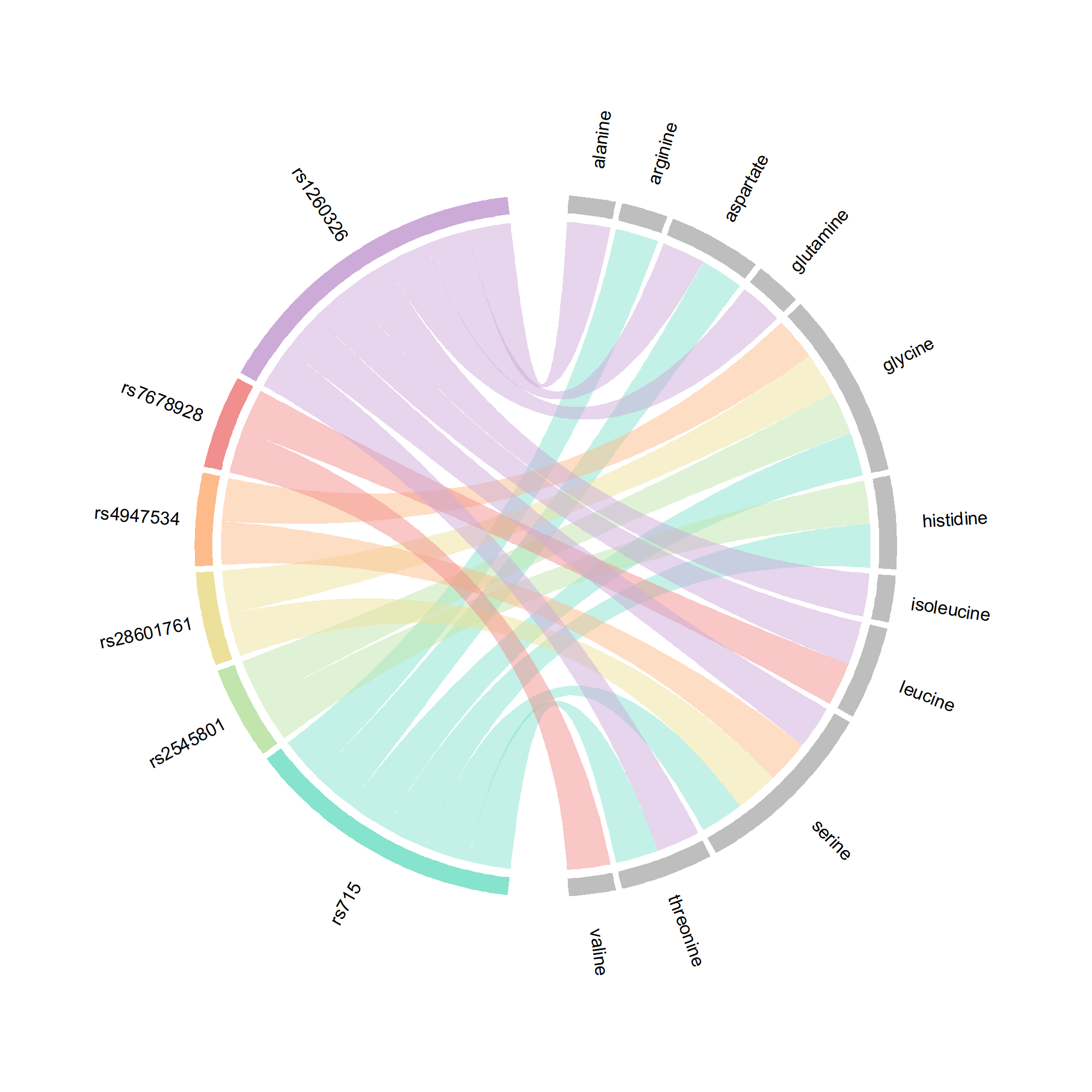

Supplementary Fig. 7. Overlap in SNPs selected as instruments for amino acids
